# The Epidemiology of Intensive Care Unit Readmissions Across Ten Health Systems

**DOI:** 10.1101/2025.03.10.25323672

**Authors:** Saki Amagai, Vaishvik Chaudhari, Kaveri Chhikara, Nicholas E. Ingraham, Chad H. Hochberg, Anna K Barker, Chengsheng Mao, Alexander C. Ortiz, Gary E. Weissman, Benjamin E. Schmid, Megan Schwinne, Sivasubramanium V. Bhavani, Shan Guleria, Zewei Liao, Nikolay Markov, Patrick G. Lyons, Brenna Park-Egan, The CLIF Consortium, William F. Parker, Yuan Luo, Juan C. Rojas, Catherine A. Gao

## Abstract

**Background:** ICU readmissions are associated with increased morbidity, mortality, and healthcare costs. As ICU patient complexity increases and care practices evolve, the contemporary epidemiology of ICU readmissions remains unclear. We aimed to examine ICU readmission rates and timing across multiple health systems, focusing on unplanned readmissions occurring within 24, 48, and 72 hours after ICU discharge.

**Methods:** We performed a retrospective cohort study using federated data from the Common Longitudinal ICU data Format (CLIF) Consortium, comprising nine healthcare systems between January 2020 and December 2021 and the MIMIC-IV database. The cohort included adult patients (≥18 years) discharged alive from the ICU. Readmissions following planned surgeries or interventional procedures were excluded. Data were analyzed locally at each site without centralizing patient-level data, and analyses focused on patient demographics, discharge disposition, readmission timing, and clinical interventions during ICU stays and readmissions. Statistical comparisons were performed using two-proportion z-tests and chi-squared tests.

**Results:** Among 185,241 hospital admissions across 19 hospitals, 8.6% of ICU discharges were readmitted during the same hospitalization. Unplanned readmissions occurred within 24 hours in 1.9% of cases, 3.4% within 48 hours, and 4.5% within 72 hours. Readmitted patients experienced higher in-hospital mortality (20.6% vs. 2.1%, p<0.001). Compared to the initial ICU stay, ICU readmissions were associated with significantly increased respiratory (42.3% vs. 35.3%, p<0.001) and vasopressor support (26.1% vs. 23.1%, p<0.001).

**Conclusions:** ICU readmissions remain common and are linked to worse outcomes. Readmissions require more respiratory and vasopressor support. Future work should focus on characterizing these subphenotypes and improving ICU discharge processes to reduce preventable readmissions.

## Background

Intensive care unit (ICU) readmissions are linked to increased patient morbidity, mortality, and healthcare costs.^1,2^ However, as patient complexity in the modern ICU increases and care practices evolve, the recent epidemiology of ICU readmissions remains unknown. Understanding the rates of ICU readmissions across multiple hospitals can inform strategies to improve patient outcomes and resource utilization. We aimed to examine ICU readmission patterns across ten health systems by analyzing both the overall occurrence of ICU readmissions during hospital stays and the timing of unplanned readmissions, focusing on those occurring within 24, 48, and 72 hours after ICU transfer.

## Methods

A retrospective cohort study was conducted using data from nine healthcare systems between January 2020 and December 2021. This work leveraged data from the Common Longitudinal ICU data Format (CLIF) Consortium,^3^ specifically: Northwestern University, Oregon Health & Science University, Rush University, Johns Hopkins University, University of Chicago, University of Michigan, University of Minnesota, Emory University, and the University of Pennsylvania. We also incorporated data from the MIMIC-IV database (Beth Israel Deaconess hospital system, 2008-2019). Adult patients (≥18 years) transferred out of the ICU alive were included, and data were analyzed on the level of each hospitalization. We did not include smaller hospitals with fewer than 100 readmissions. Data collected encompassed patient demographics, discharge disposition (for example, home vs died), and readmission status. An unplanned ICU readmission was defined as a transfer from the ICU to the regular hospital floor and then back to the ICU during the same hospitalization. If a patient had multiple ICU readmissions during their hospitalization, we examined only the first ICU readmission. If a patient was transferred to another hospital during the same hospitalization, we combined these encounters and treated them as a single continuous hospitalization. ICU readmissions following surgical or interventional procedures were not included in the readmitted cohort, with the assumption that these readmissions were potentially planned by the clinical service before the procedure. We evaluated patterns of hemodynamic support (the use of vasopressors norepinephrine, epinephrine, phenylephrine, vasopressin, angiotensin) and respiratory support [including invasive mechanical ventilation (IMV), non-invasive positive pressure ventilation (NIPPV), and high-flow nasal cannula (HFNC)]. We compared requirements during initial ICU stays versus readmissions, to identify clinical triggers for critical care readmission. We used a two-proportion z-test to compare respiratory support rates at ICU admission and readmission, and a chi-squared test to assess differences between readmitted and non-readmitted patients. LLMs were used to assist coding and drafting, with all output reviewed by authors, who take responsibility for the resulting data. The full code for our analysis is available at: https://github.com/Common-Longitudinal-ICU-data-Format/CLIF_icu_readmission. The reproducibility of the code has been demonstrated as each site ran the code locally within their own secure servers. The code can be fully reproduced using the CLIF-MIMIC pipeline to format MIMIC-IV into CLIF format and running the ICU readmission analysis.

## Results

The study cohort comprised 185,241 total hospital admissions (mean age 61.3, 44.7% female, 23.7% African American, 5.5% Hispanic) from 19 hospitals (Supplemental Table 1 for full demographics). An example Sankey flow diagram of ICU readmissions from an academic hospital is shown in **Figure 1A**. Among these ICU discharges, 8.6% (range: 4.7-12.9%) were readmitted during the same hospitalization, with 1.9% (range: 1.1-2.9%) occurring within 24 hours, 3.4% (range: 1.9-4.5%) within 48 hours, 4.5% (range: 2.5-5.6%) within 72 hours of ICU discharge (**Figure 1B**). Across all healthcare systems, readmitted patients exhibited a higher in-hospital mortality rate compared to non-readmitted patients (2.1% vs 20.6%, p<0.001) (**Figure 1C**). Hospitals with step-down units had a 7.0% readmission rate (range: 4.6-8.3%). Analysis of clinical interventions revealed significantly higher rates of respiratory support (35.3% vs 42.3%, p<0.001) and vasopressor support (23.1% vs 26.1%, p<0.001) during ICU readmissions compared to index ICU stays.

**Figure 1.**
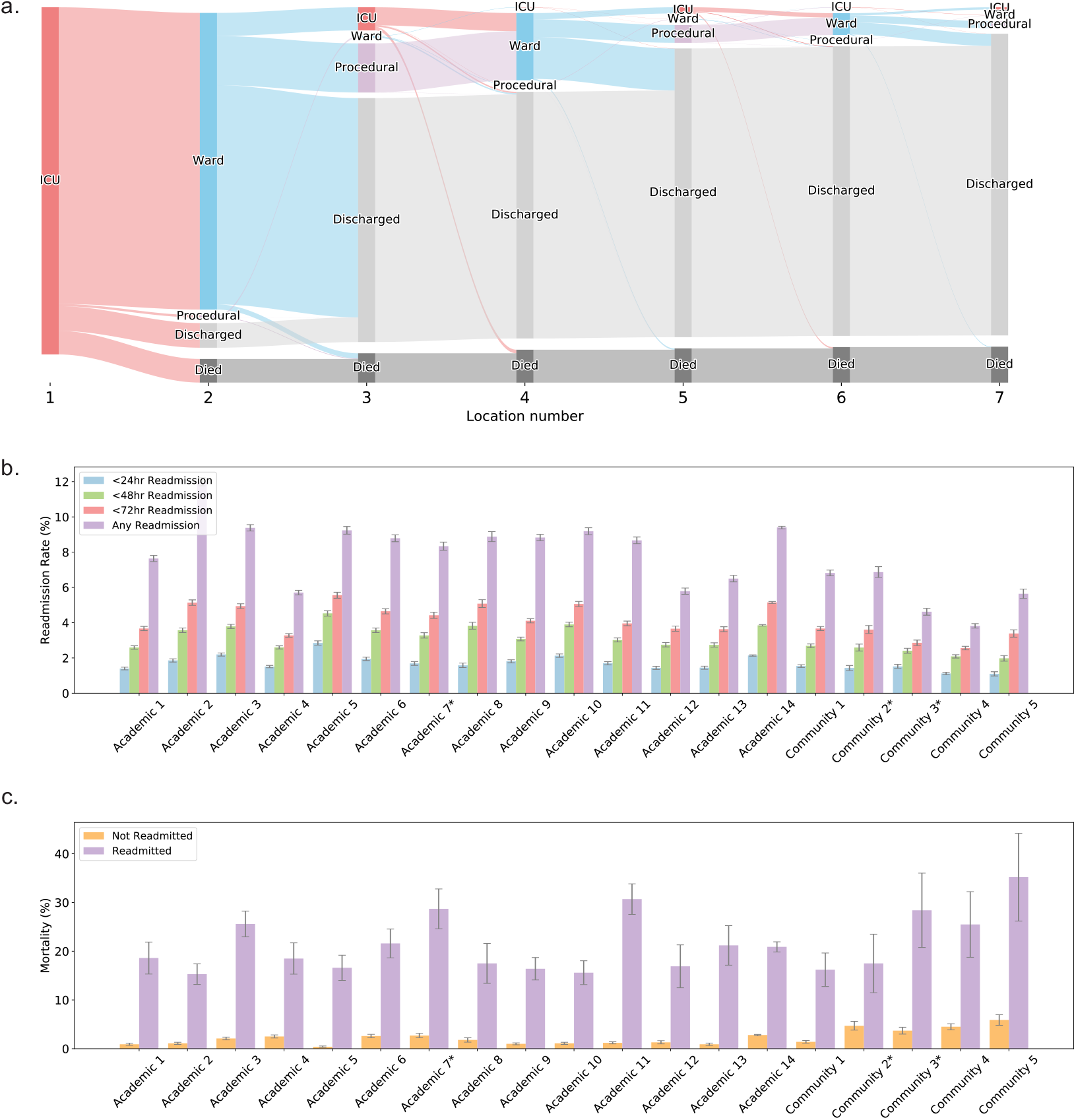
Multi-site comparison of ICU readmission rates. An unplanned ICU readmission was defined as a transfer from the ICU to a regular hospital floor and then back to the ICU within the same hospitalization. **a**. Example Sankey diagram from Academic Hospital 3 illustrating the patient journey within the hospital after index ICU admission. **b**. Comparison of readmission rates across hospitals at different time thresholds (<24 hr, <48 hr, <72 hr, and any readmission). **c**. Comparison of mortality rates between readmitted (at any time during hospitalization) and non-readmitted patients across academic and community hospitals. * indicates a hospital with a stepdown unit. The 95% confidence interval was estimated using the Wald method for binomial success probability, representing the range within which we are 95% confident that the true values of the observed trend data points lie.

**Figure 2.**
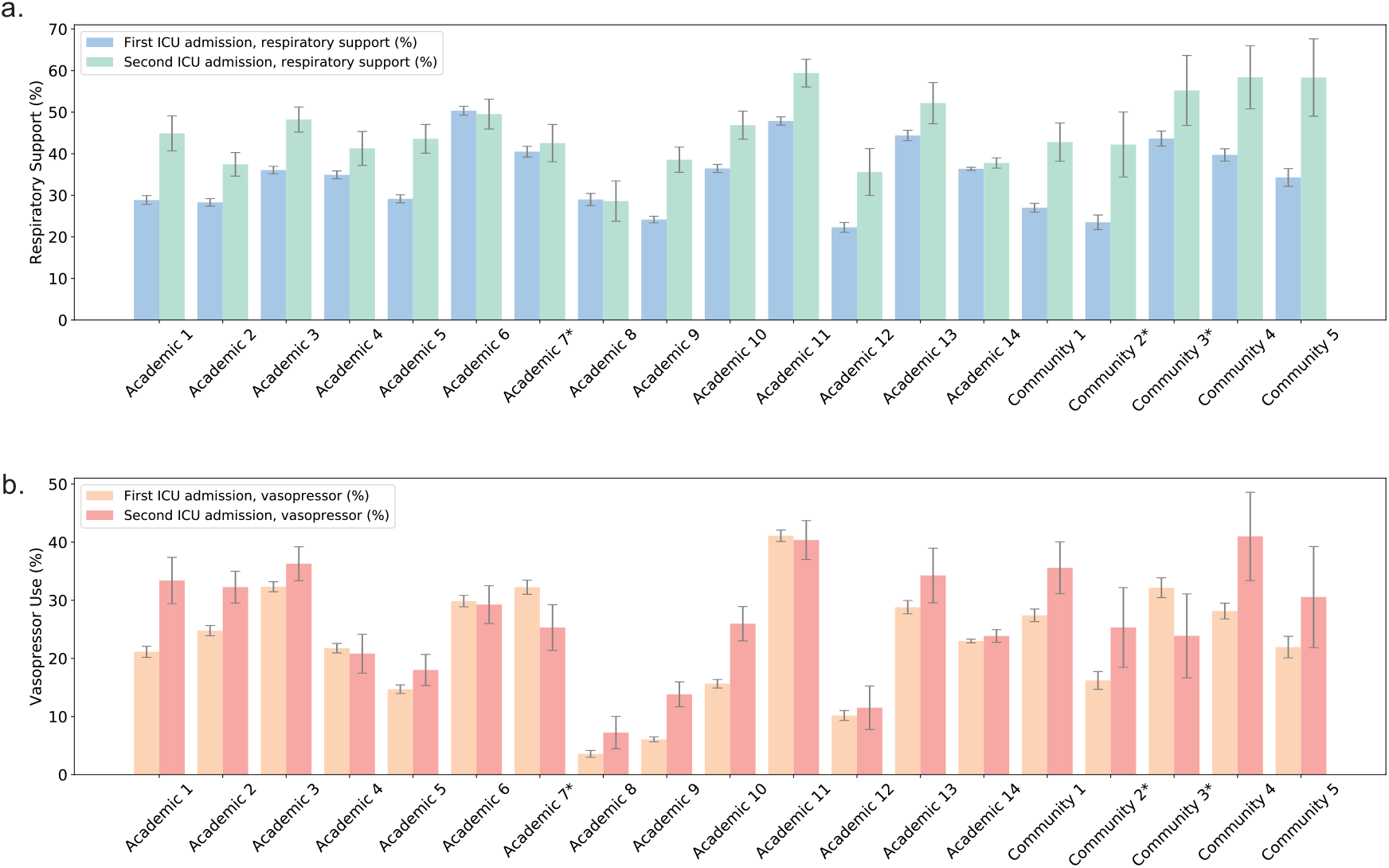
Patterns of hemodynamic and respiratory support interventions were compared between initial ICU stays versus readmissions. **a**. Comparison of respiratory support (IMV, NIPPV, HFNC) usage between the first ICU admission and subsequent readmission. **b**. Comparison of vasopressor (norepinephrine, epinephrine, phenylephrine, vasopressin, angiotensin) usage between the first ICU admission and subsequent readmission. The 95% confidence interval was estimated using the Wald method for binomial success probability, representing the range within which we are 95% confident that the true values of the observed trend data points lie. * indicates a hospital with a stepdown unit.

## Discussion

Our study underscores the continued prevalence of ICU readmissions across multiple hospitals and its associated higher mortality. We examined shock and respiratory failure rates upon readmission, and future work should focus on characterizing these subphenotypes. There was a trend toward lower ICU readmission rates in hospitals with stepdown units; however, it is challenging to disentangle the specific contributing factors given variations in healthcare systems and policies. We demonstrated the flexibility of our federated consortium to examine clinical questions across multiple sites. Identifying high-risk patients and potentially modifiable factors before ICU transfer are crucial next steps.^4^ Future work should focus on enhanced transfer planning,^5,6^ and post-ICU monitoring strategies to optimize readmission rates and improve patient outcomes.

## Supporting information

Supplemental: CLIF Consortium Author List

## Data Availability

The data generated in this study are not publicly available, as they contain information that could compromise the privacy of research participants.

## Data availability statement

The underlying data from each institution are not publicly available due to privacy, regulatory, and institutional restrictions. This study was conducted using a federated analysis approach, without centralizing patient level data at any single site. Data were analyzed locally at participating institutions, ensuring that patient-level data remained within each contributing site. The CLIF Consortium welcomes opportunities for collaboration and data sharing through a standard project intake and approval process, which includes review and approval by the consortium’s leadership and participating sites. Researchers interested in collaborating with the CLIF Consortium are encouraged to contact the consortium for more information.

## Code availability

The full code for our analysis is available at: https://github.com/Common-Longitudinal-ICU-data-Format/CLIF_icu_readmission

## Funding

SA is supported by AHA 25PRE1372516.

NI is supported by NIH/NHLBI K23HL166783.

CHH is supported by NIH/NHLBI K23HL169743.

AB is supported by an institutional research training grant (NIH/T32-HL 00774).

ACO is supported by an institutional research training grant (NIH/T32-HL-007891).

GEW received support from R35GM155262 relevant to the published work.

MS is supposed by NIH UL1TR002378.

SVB is supported by NIH/NIGMS K23GM144867.

NM is supported by AHA 24PRE1196998

PGL is supported by NIH K08CA270383.

WFP is supported by NIH K08HL150291, R01LM014263, and the Greenwall Foundation.

YL is supported in part by NIH U01TR003528 and R01LM013337.

JCR is supported by NIH/NIDA R01DA051464 and the Searle Funds at The Chicago Community Trust.

CAG is supported by NIH/NHLBI K23HL169815, a Parker B. Francis Opportunity Award, and an American Thoracic Society Unrestricted Grant.

## COI

JCR discloses consulting fees from Truveta. Other authors declare no commercial conflicts of interest.

## List of sites and IRBs

Emory University: Study 1815

Northwestern University: STU00202840

Oregon Health & Science University: 00025188

Rush University: IRB 240828040

The Johns Hopkins Health System Corporation: IRB00421735

University of Chicago: IRB20-1823

University of Michigan: HUM00260807

University of Minnesota: STUDY00014815

University of Pennsylvania: 853717

MIMIC: publicly-available, deidentified database available to credentialed users having signed a DUA.

## Supplementary Materials

**Supplemental Table 1.** Aggregated admission characteristics and outcomes by readmission status. All variables across sites were pooled to derive the overall sample characteristics. The p-values were obtained from chi-squared tests.

|  | Not readmitted<br>(n=169,395) | Readmitted<br>(n=15,846) | P-value |
| --- | --- | --- | --- |
| Age, mean (SD) | 61.1 (17.2) | 63.1 (15.8) | <0.001 |
| ICU length of stay, days, mean (SD) | 3.6 (5.8) | 5.7 (9.8) | <0.001 |
| ICU readmission, hours, mean (SD) | NA | 123.1 (205.0) | NA |
| Sex: Female, n (%) | 75973 (44.8%) | 6756 (42.6%) | <0.001 |
| Race: Black, n (%) | 40709 (24.0%) | 4086 (25.3%) | <0.001 |
| Race: White, n (%) | 103882(61.3%) | 9395 (59.3%) |  |
| Race: Asian, n (%) | 5760 (3.4%) | 547(3.5%) |  |
| Race: Other/Unknown, n (%) | 19044 (11.2%) | 1818 (11.5%) |  |
| Ethnicity: Hispanic, n (%) | 9263 (5.5%) | 815 (5.1%) | <0.001 |
| In-hospital Mortality, n (%) | 3593 (2.1%) | 3257 (20.6%) | <0.001 |

**Supplemental Table 2.** Hospital labels by health system.

| Health System | Hospital Labels |
| --- | --- |
| Health System 1 | Academic 1, Community 1, Academic 2 |
| Health System 2 | Community 2*, Academic 3, |
| Health System 3 | Academic 4 |
| Health System 4 | Academic 5 |
| Health System 5 | Academic 6 |
| Health System 6 | Community 3*, Community 4, Academic 7* |
| Health System 7 | Academic 8, Academic 9, Community 5 |
| Health System 8 | Academic 10 |
| Health System 9 | Academic 11, Academic 12, Academic 13 |
| Health System 10 | Academic 14 |

**Supplemental Figure 1.**
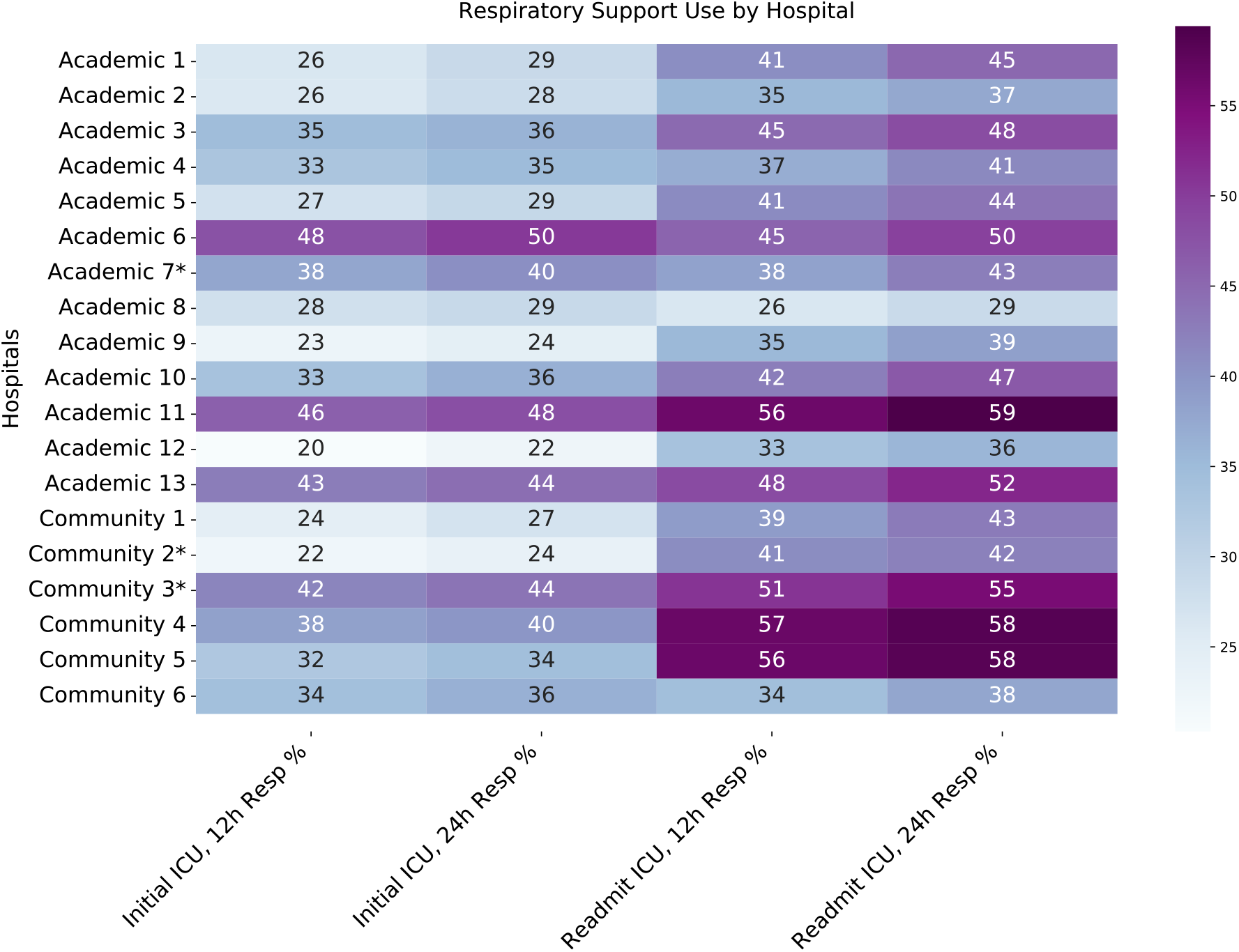
Heat map for respiratory support (IMV, NIPPV, HFNC) usage, split by first ICU admission versus readmission, by hours after readmission requiring support. * indicates a hospital with a stepdown unit.

**Supplemental Figure 2.**
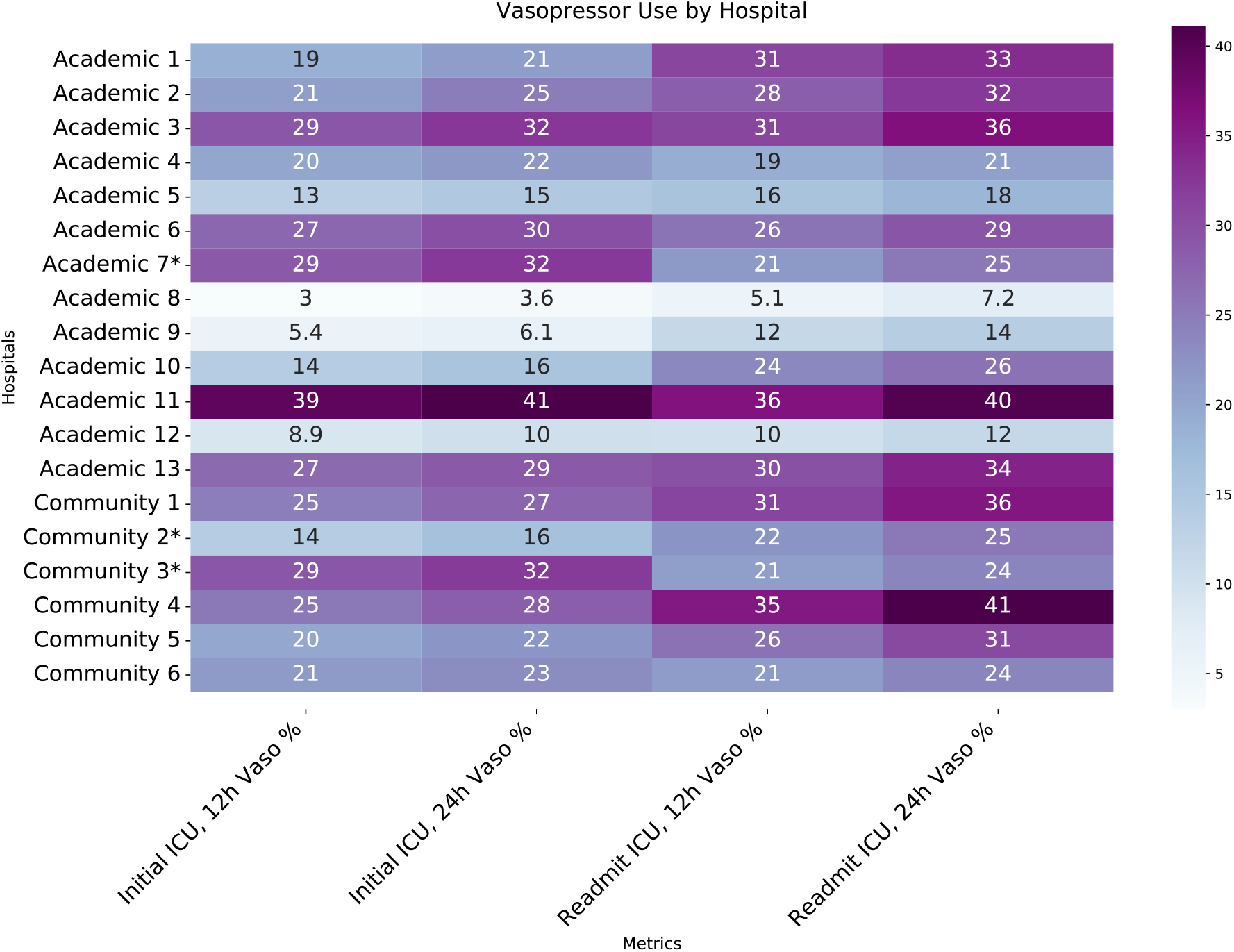
Heat map for vasopressor (norepinephrine, epinephrine, phenylephrine, vasopressin, angiotensin) usage, split by first ICU admission versus readmission, by hours after readmission requiring support. * indicates a hospital with a stepdown unit.

## Notes

### Competing Interest Statement

The authors have declared no competing interest.

### Author Declarations

Ethical approval for this work was granted by the following ethics committees/IRBs: The Institutional Review Board of Oregon Health & Science University gave ethical approval for this work (IRB approval number: 00025188). The Institutional Review Board of the University of Minnesota gave ethical approval for this work (IRB approval number: STUDY00014815). The Institutional Review Board of the University of Michigan gave ethical approval for this work (IRB approval number: HUM00144238). The Institutional Review Board of Northwestern Medicine gave ethical approval for this work (IRB approval number: STU00202840). The Institutional Review Board of the University of Chicago gave ethical approval for this work (IRB approval number: IRB20-1823). The Institutional Review Board of Rush University gave ethical approval for this work (IRB approval number: 20082408-IRB01). The Institutional Review Board of The Johns Hopkins Health System Corporation gave ethical approval for this work (IRB approval number: IRB00421735). The Institutional Review Board of Emory University gave ethical approval for this work (IRB approval number: Study 1815). The Institutional Review Board of the University of Pennsylvania gave ethical approval for this work (IRB approval number: 853717). Each site had waiver of consent.

